# Fetal sexual dimorphism and preeclampsia among twin pregnancies

**DOI:** 10.1101/2023.11.10.23298403

**Authors:** Rebekah E. Brown, Akaninyene I. Noah, Ashley V. Hill, Brandie DePaoli Taylor

## Abstract

**Background:** In singleton pregnancies, fetal sexual dimorphism has been observed in hypertensive disorders of pregnancy (HDP), particularly preeclampsia, a morbid syndrome that increases risk of adult onset cardiovascular disease for mothers and their offspring. However, few studies have explored the effect of fetal sex on HDP among twin pregnancies.

**Methods:** We conducted a retrospective cohort study of 1,032 twin pregnancies between 2011 - 2022 using data from a perinatal database that recruits participants from three hospitals in Houston, TX. We categorized pregnancies based on fetal sex pairings into female/female, male/male, and female/male. Pregnancies with a female/female fetal sex were used as our reference group. Our primary outcomes included gestational hypertension, preeclampsia, superimposed preeclampsia, and preeclampsia subtyped by gestational age of delivery. A modified Poisson regression model with robust error variance was used to calculate the relative risk (RR) and 95% confidence interval (CI) for the association between fetal sex pairs and HDP.

**Results:** Adjusted models of female/male fetal sex pairs were associated with preterm preeclampsia (RR 2.01, 95% CI 1.15-3.53) relative to those with female/female fetuses. No associations with other HDP were observed among pregnancies with male/male fetal sex compared to those with female/female fetal sex pairs.

**Conclusions:** We found some evidence of sexual dimorphism for preterm preeclampsia among female/male twin pairs. Additional research is needed to understand what biological mechanisms could explain these findings.

## Introduction

One in 32 pregnancies in the United States are multifetal^1^ which increases the risk of adverse perinatal outcomes including hypertensive disorders of pregnancy (HDP), preterm birth, small for gestational age, and other complications.^2,3^ In the case of preeclampsia, a morbid and deadly HDP, multifetal pregnancy increases risk of early onset preeclampsia and preeclampsia with severe features,^2^ which may be due to elevated cardiovascular burden.^4^ This is a significant public health concern. Preeclampsia affects 5-10% of pregnancies^5^ but the pathogenesis is not fully understood, and the only treatment is delivery, which often results in preterm birth.^6^ Furthermore, women with preeclampsia have a 3-fold increased risk of cardiovascular disease^6^ and their infants also have increased risk of cardiovascular disease later in life.^7,8^ Given the serious nature of preeclampsia, and the long-term impacts on cardiovascular health, further research on preeclampsia in multifetal pregnancies is warranted.

In singleton pregnancies, the sex of the fetus is thought to impact the risk of preeclampsia, possibly through differences in placental metabolic and immunological function or through maternal systemic angiogenic and cytokine profiles.^9–14^ This suggests, that male and female fetuses may respond differently to maternal physiological responses and other underlying stimuli. In multifetal pregnancies, the physiologic changes that occur are essentially an amplified version of a singleton pregnancy.^15^ But few studies have explored if fetal sex alters risk of preeclampsia or other HDP in twin pregnancies.

Although data is limited, studies in multifetal pregnancies align with trends seen in singleton pregnancies with male sex being associated with shorter gestation and male/male pregnancies at the highest risk for preterm labor.^16,17^ Approximately three studies have examined fetal sex and preeclampsia in multifetal pregnancies. These studies were carried out in Japan, Sweden, and Slovenia,^3,17,18^ but results were inconsistent, and only one of the three studies accounted for preeclampsia subtypes which are thought to have different pathophysiologies.^6^ For example, preeclampsia can be subtyped by early and late onset, where gestational age of delivery is often used as a proxy (i.e. term and preterm preeclampsia). Defective trophoblast invasion and spiral artery remodeling is consistently linked to early onset preeclampsia.^19^ Those with early onset preeclampsia are also more likely to have growth restricted fetuses and have higher risk of cardiovascular outcomes.^6^ In studies of fetal sex in singletons there are differences observed in the association between fetal sex based on subtypes of preeclampsia.^10^ Male fetal sex has been associated with preeclampsia but there appears to be a female bias in preeclampsia that occurs preterm among singletons.^10,20,21^ This suggests that that in singletons fetal sex may influence severity of preeclampsia. Given the limited data on the impact of fetal sex on HDP in twin pregnancies, our objective was to examine the association between fetal sex and various HDP including preeclampsia subtypes among twin pregnancies.

## Methods

### Study Design

We conducted a retrospective cohort study of 1,032 twin deliveries using data from Peribank^22^, an obstetrical database affiliated with Baylor College of Medicine and Texas Children’s Hospital. Peribank contains information pertaining to pregnancy, delivery, and the postnatal course. Pregnant individuals are recruited when they present to labor and delivery or immediately after delivery in cases of emergency.^22^ Data is obtained through patient interviews and electronic medical records. To be included, participants must be pregnant, ≥ 18 years of age (or ≥16 years of age if emancipated), able to read and understand the consent form, and able to sign the consent form.^22^ There are no exclusion criteria. The Institutional Review Boards at Baylor College of Medicine and Texas Children’s Hospital approved Peribank. Informed consent was obtained from each participant. For our study, the Institutional Review Board at the University of Texas Medical Branch determined that this analysis was exempt. One author had full access to all the data in the Peribank database and takes full responsibility for its integrity and data analysis.

### Primary Exposure

Our primary exposure was multifetal pregnancy with female/female, male/male, or female/male fetal sex pairs as determined in the administrative database. Approximately 18 women were excluded due to missing data on fetal sex (1.74%), the remaining 1,014 participants were included in our analysis.

### Primary Outcome

The primary outcomes were hypertensive disorders of pregnancy (HDP) which included preeclampsia, gestational hypertension, and superimposed preeclampsia diagnosed using the American College of Obstetricians and Gynecologists (ACOG) criteria.^23^ Preeclampsia is diagnosed by “systolic blood pressure of 140 mm Hg or more or diastolic blood pressure of 90 mm Hg or more on two occasions at least 4 hours apart after 20 weeks of gestation in a woman with a previously normal blood pressure” and proteinuria “at least 300 mg per 24-hour urine collection or protein/creatinine ratio of 0.3 mg/dL” or evidence of systemic organ dysfunction in the absence of protienuria.^23^ In addition, preeclampsia can be defined based on gestational age of delivery and whether preeclampsia results in preterm birth <37 weeks gestation (i.e. preterm preeclampsia and term preeclampsia). Gestational hypertension is defined as hypertension without proteinuria or severe features occurring after 20 weeks gestation with blood pressure levels returning to normal in the postpartum period.^23^ Superimposed preeclampsia is defined as preeclampsia superimposed upon previously diagnosed chronic hypertension.^23^

### Covariates

Information was collected regarding the age of the mother (median, IQR), body mass index (BMI), racial/ethnic background (Non-Hispanic (NH) White, Hispanic, NH Asian/Other), foreign-born status (no, yes), marital status (married, single), education level (some college and above, high school, and less than high school), and insured status (private, Medicaid/CHIP, other/unknown/no insurance). Data on alcohol (no, yes), tobacco (no, yes), and drug use (no, yes) was obtained. Information on maternal comorbidities (endometriosis, chronic health conditions, thyroid disease, mental health issues, seizure disorder, sickle cell disease, hypospadias, prior gestational diabetes) (no, yes) was collected. Data was also available on the number of previous pregnancies (no pregnancy, 1-2 prior, >2 prior pregnancies) and the gestational age at first prenatal visit by trimester were collected (0-12 weeks, >12 weeks).

### Statistical Analyses

We compared maternal demographic and clinical characteristics between fetal sex pairs (female/female, male/male, and female/male) using a modified Poisson regression model with robust error variance, which is an accepted method to directly measure relative risk (RR) and 95% confidence intervals (CI) risk rather than using odds ratios as estimates for risk.^24^ The same modeling approach was used to examine fetal sex pairs and risk of HDP. Pregnancies with a female/female fetal sex pair were used as our reference group as male fetal sex is most consistently linked with elevated risks of adverse pregnancy outcomes.^16,17^ Our model covariates were selected a priori. They included maternal age, maternal comorbid status (yes/no), education level, and insurance status. The missingness among the variables included in our model ranged from 1 - 6%. We used fully conditional multiple imputation with ten iterations to address missing data. All analyses were conducted using SAS version 9.4, Cary, North Carolina.

## Results

### Population and Characteristics

In our study population there were a total of 1,014 women with multifetal pregnancies, including 337 (33.2%) with female/female, 345 (34.0%) with male/male, and 332 (32.7%) with female/male pairs (**Table 1**). Approximately 40.5% of women were foreign-born, while 57.0% were US born. Foreign-born mothers were less likely to have female/male twins compared to female/female (RR 0.77, 95% CI 0.62-0.97). Mothers with a high school education or less (RR 0.65, 95% CI 0.47-0.91) and those insured through Medicaid/CHIP (RR 0.78, 95% CI 0.63-0.97) were less likely to have female/male pairs compared to female/female. Mothers of female/female, male/male, and female/male twin pairs had a median BMI of 32.9, 31.3, and 32.4, respectively. Median BMI was slightly lower in mothers of male/male twin pairs compared to those with female/female (RR 0.97, 95% CI 0.96-0.99).

**Table 1.** Maternal demographics by fetal sex pairs.

| **Table 1. Maternal demographics by fetal sex pairs** |  |  |  |  |  |
| --- | --- | --- | --- | --- | --- |
| Variables | Female/Female | Male/Male | *Risk Ratio (RR)<br>RR, 95% CI | Female/Male | *Risk Ratio (RR)<br>RR, 95% CI |
| N | 337 (33.2) | 345 (34.0) |  | 332 (32.7) |  |
| *Age (Median, IQR)* | 32 (27 – 25) | 31 (27 – 35) | 0.99, 0.98 – 1.01 | 32 (28 – 35) | 1.01, 1.00 – 1.02 |
| *Race/Ethnicity, (n, %)* |  |  |  |  |  |
| Non-Hispanic White | 97 (28.4) | 101 (29.2) | Ref | 105 (31.7) | Ref |
| Non-Hispanic Black | 53 (15.5) | 57 (16.5) | 1.02, 0.73 - 1.41 | 85 (25.7) | 1.18, 0.89 – 1.58 |
| Hispanic | 169 (49.4) | 163 (47.1) | 0.96, 0.75 – 1.23 | 118 (35.7) | 0.79, 0.61 – 1.03 |
| NH Asian/Other | 23 (6.7) | 25 (7.3) | 1.02, 0.66 – 1.58 | 23 (6.9) | 0.96, 0.61 – 1.51 |
| *Foreign-born Status (n, %)* |  |  |  |  |  |
| No | 181 (54.4) | 190 (55.6) | Ref | 207 (65.9) | Ref |
| Yes | 152 (45.7) | 152 (44.4) | 0.98, 0.79 – 1.21 | 107 (34.1) | 0.77, 0.62 – 0.97 |
| *Marital Status (n, %)* |  |  |  |  |  |
| Married | 264 (77.9) | 270 (78.3) | Ref | 264 (79.8) | Ref |
| Single | 75 (22.1) | 75 (21.7) | 1.0, 0.77 – 1.29 | 67 (20.2) | 0.95, 0.73 – 1.24 |
| ***Education, (n, %)*** |  |  |  |  |  |
| Some College and above | 186 (57.6) | 197 (60.4) | Ref | 214 (69.5) | Ref |
| High School | 66 (20.4) | 79 (24.2) | 1.06, 0.82 - 1.36 | 57 (18.5) | 0.87, 0.66 - 1.15 |
| Less than High School | 71 (22.0) | 50 (15.3) | 0.81, 0.60 - 1.09 | 37 (12.0) | 0.65, 0.47 - 0.91 |
| ***Insurance (n, %)*** |  |  |  |  |  |
| Private | 134 (40.2) | 143 (41.7) | Ref | 170 (52.0) | Ref |
| Medicaid / CHIP | 189 (56.8) | 188 (54.8) | 0.98, 0.79 - 1.21 | 145 (44.2) | 0.78, 0.63 - 0.97 |
| Other / Unknown / No Insurance | 10 (3.1) | 12 (3.7) | 1.06, 0.59 - 1.92 | 12 (3.7) | 0.98, 0.54 - 1.75 |
| ***Alcohol Use, n (%)*** |  |  |  |  |  |
| No | 151 (44.2) | 153 (44.2) | Ref | 149 (45.2) | Ref |
| Yes | 191 (55.9) | 192 (55.8) | 0.99, 0.81 - 1.23 | 182 (54.9) | 0.98, 0.79 - 1.22 |
| ***Smoking, (n, %)*** |  |  |  |  |  |
| No | 285 (83.3) | 295 (85.3) | Ref | 275 (83.3) | Ref |
| Yes | 57 (16.7) | 51 (14.7) | 0.93, 0.70 - 1.25 | 55 (16.7) | 1.0, 0.75 - 1.34 |
| ***Drug Use, (n, %)*** |  |  |  |  |  |
| No | 305 (90.5) | 313 (90.7) | Ref | 305 (92.2) | Ref |
| Yes | 32 (9.5) | 32 (9.3) | 0.99, 0.76 - 1.28 | 26 (7.9) | 0.90, 0.67 - 1.21 |
| ***Body Mass Index (BMI),***<br>***Median (IQR)*** | 32.9,<br>(29.8–37.3) | 31.3<br>(28.1–35.5) | 0.97, 0.96–0.99 | 32.4<br>(28.7–37.1) | 0.99, 0.98 – 1.01 |
| ***Maternal Comorbidities*** |  |  |  |  |  |
| No | 111 (32.7) | 108 (31.3) | Ref | 79 (24.0) | Ref |
| Yes | 229 (67.4) | 237 (68.7) | 1.03, 0.82 - 1.29 | 250 (76.0) | 1.25, 0.97 - 1.61 |
| ***Gravidity (n, %)*** |  |  |  |  |  |
| No prior pregnancy | 104 (30.9) | 104 (30.2) | Ref | 88 (26.6) | Ref |
| 1 – 2 prior pregnancies | 135 (40.1) | 161 (46.8) | 1.08, 0.85 - 1.39 | 143 (43.2) | 1.12, 0.86 - 1.46 |
| > 2 prior pregnancies | 98 (29.1) | 79 (23.0) | 0.88, 0.66 - 1.18 | 100 (30.2) | 1.07, 0.81 - 1.43 |
| ***Gestational Age at 1<sup>st</sup> visit*** |  |  |  |  |  |
| 0 – 12 weeks | 194 (64.7) | 195 (64.1) | Ref | 208 (69.1) | Ref |
| >12 weeks | 106 (35.3) | 109 (35.9) | 1.01, 0.81 - 1.25 | 93 (30.9) | 0.89, 0.71 - 1.13 |
| *Risk ratio was calculated using a modified Poisson regression model with robust estimates. |  |  |  |  |  |

We examined the association between fetal sex and hypertensive disorders in multifetal pregnancies. Within our cohort, approximately 104 (10.8%) were diagnosed with preeclampsia, 35 (3.4%) were diagnosed with gestational hypertension, and 24 (2.7%) were diagnosed with superimposed preeclampsia (**Table 2**). Participants carrying female/male twins were more likely to have preeclampsia compared to female/female (RR 1.71, 95% CI 1.04-2.81) but this was not observed for male/male pairs (RR 1.45, 95% CI 0.88-2.41). There was no association between male/male (RR 0.99, 95% CI 0.41-2.38) and female/male (RR 1.50, 95% CI 0.66-3.42) pairs and gestational hypertension when compared to female/female. Similar to gestational hypertension, there was no difference in the risk of superimposed preeclampsia between male/male (RR 2.06, 95% CI 0.70-6.07) and female/male (RR 1.80, 95% CI 0.60-5.44) compared to female/female twin pairs.

**Table 2.**
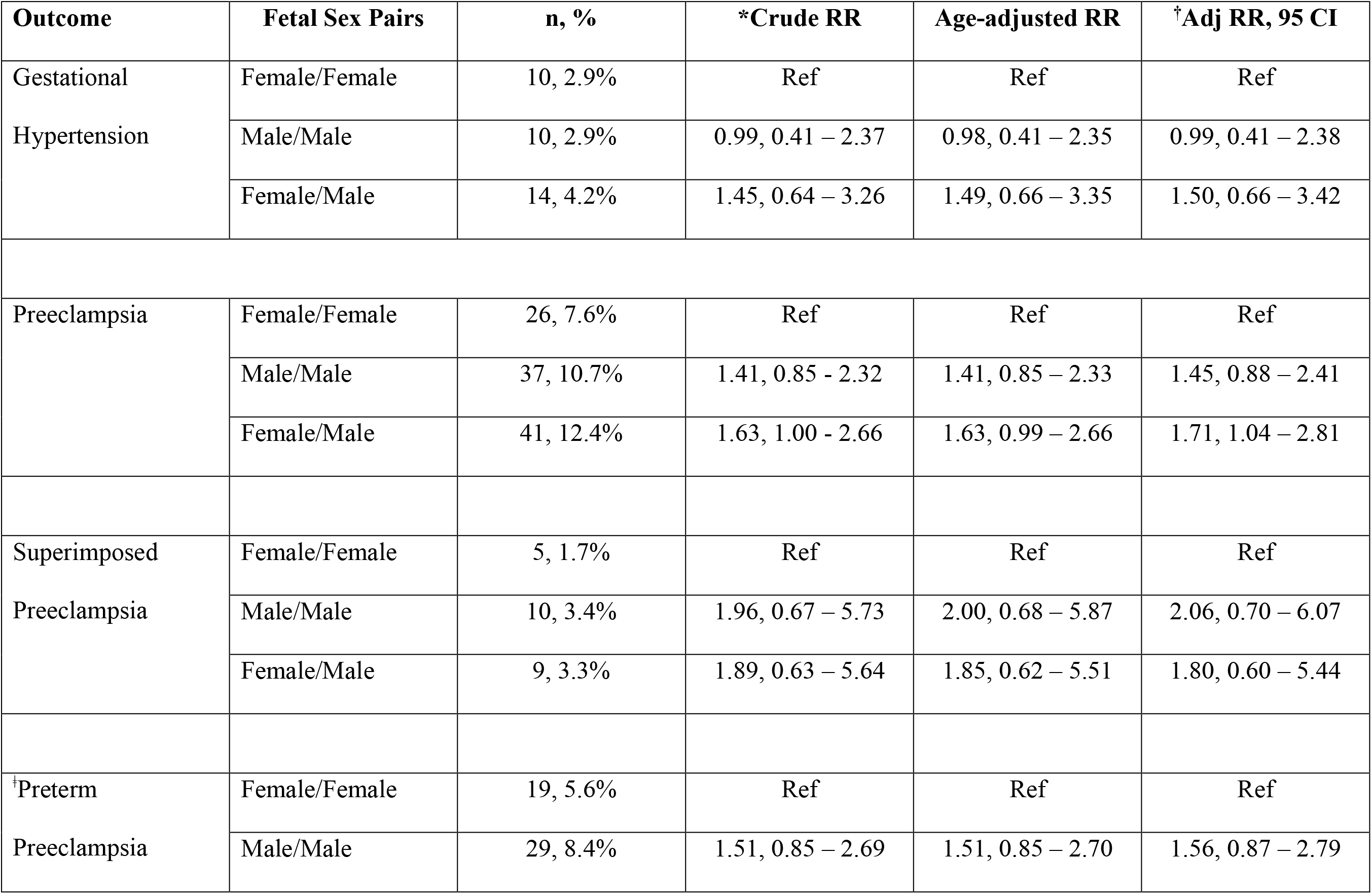
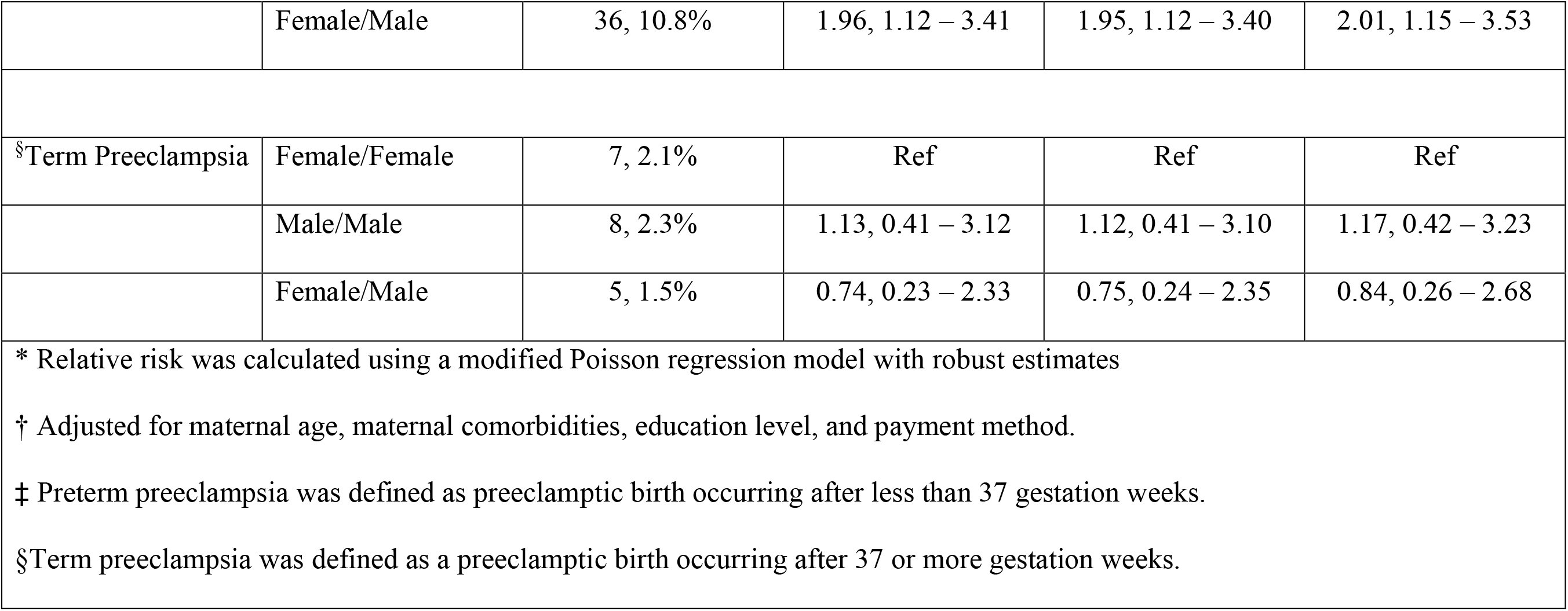
Association between fetal sex pairs and hypertensive disorders of pregnancy.

Of those with preeclampsia, 84 (8.1%) were diagnosed with preterm preeclampsia while 20 (1.9%) were diagnosed with term preeclampsia (**Table 2**). Female/male pairs were more likely to have preterm preeclampsia compared to female/female (RR 2.01, 95% CI 1.15-3.53), while no difference was noted in male/male (RR 1.45, 95% CI 0.88-2.41). There was no difference in term preeclampsia for the male/male (RR 1.56, 95% CI 0.42-3.23) and female/male (RR 0.84, 95% CI 0.26-2.68) pairs compared to female/female.

Lastly, as a sensitivity analysis, to be consistent with one of the previous papers examining twin fetal sex and preeclampsia, we explored if associations between female/male pairs were consistent if male/male were the reference group. We found no association between female/male fetal sex pairs and preeclampsia (RR 1.17, 95% CI 0.75-1.83), using male/male pairs as our reference groups. However, effect estimates were in a similar direction as our analysis using female/female twins as the reference group. We also found no association between female/male pairs and preterm preeclampsia (RR 1.29, 95% CI 0.79-2.11) compared to male/male twins, although effect estimates were also in the same direction. No associations were found between female/male twins and preeclampsia, gestational hypertension, and superimposed preeclampsia, relative to male/male fetal sex pairs.

## Discussion

We observed that individuals carrying female/male fetal sex pairs were more likely to have preterm preeclampsia compared to those carrying female/female pairs. There was no difference in preeclampsia risk among male/male twin pairs compared to female/female although effect estimates were in a similar direction. We did not observe any difference in preterm preeclampsia for female/male fetal sex pairs, when using male/male twins as our reference, although the effect estimates were in the same direction. Steen et al.,^17^ in 16,045 twin pregnancies in Sweden, found that preterm preeclampsia was more common in pregnant individuals carrying female/male or female/female twin pairs compared to those with male/male. Other studies have described differing results.^3,18^ Funaki et al., among a Japanese population, found that male/male pairs have higher risk of preterm birth but lower risk of preeclampsia when compared to female/female fetuses, although associations were only significant among dichorionic twins.^3^ Relative risks were similar for preeclampsia when they examined male/female dichorionic twin pairs but confidence intervals overlapped one.^3^ In contrast, Lučovnik et al., in a study conducted in Slovenia found no associations between monozygotic fetal sex pairs and preeclampsia compared to female/male dizygotic pairs, suggesting no influence of zygosity.^18^ Overall, prior studies are conflicting but of the handful of studies conducted only one considered subtypes of preeclampsia. Our study suggests that unlike twin pairs may be associated with preterm preeclampsia. Steen et al., distinguished between subtypes and observed an increased risk of preterm preeclampsia in women carrying female/male twin pairs but compared to male/male twin pairs.^17^ In contrast, we observed no significant difference in preterm preeclampsia risk for female/male twins compared to male/male pairs, although the effect estimates were in a similar direction.

The pathogenesis of preeclampsia is not fully understood and complicated by its heterogeneous nature. Preeclampsia consists of varying subtypes (e.g., preterm vs. term, with or without severe features,.) each hypothesized to have distinct pathogenic mechanisms, however, there are no biomarkers or clinical indicators that can distinguish subtypes.^6,19,25^ Preterm preeclampsia is hypothesized to occur in the majority of cases due to abnormal placentation caused by failed spiral artery remodeling, leading to the development of placental ischemia.^26^ In contrast, term preeclampsia may be a consequence of maternal factors such as obesity, or diabetes.^27^ According to Roberts et al., the placenta is likely normal, and preeclampsia in late pregnancy may be precipitated by a combination of pregnancy-induced physiological stress and preexisting maternal health conditions.^6^ Preeclampsia subtypes also have varying risks of long-term cardiovascular complications, with those with early onset preeclampsia having the highest risk of cardiovascular disease.^19^ However, twin pregnancies are not frequently included in preeclampsia studies and Bergman et al^28^ recently found that cardiovascular risk patterns in multifetal pregnancies with preeclampsia do not mimic singleton pregnancies. The authors suggested that perhaps preeclampsia is driven by different mechanisms in multifetal pregnancies. In our study, our findings also did not closely mimic findings from singleton pregnancies where female fetal sex is consistently linked to preterm preeclampsia.^10,20,29^

Although the impact of fetal sex on maternal outcomes is well documented, the mechanisms explaining these associations are not entirely clear. Male and female fetuses can both impact the maternal immune system in distinct ways, therefore, immunological factors may provide a link between fetal sex and preeclampsia. The role of the immune system in preeclampsia has been previously described.^30^ Female fetal sex is associated with lower first trimester pro-inflammatory markers (IFNγ and IL-12) and increased second trimester pro-inflammatory (TNFβ and IL1β), anti-inflammatory (IL4r), and regulatory cytokines (IL5 and IL10).^10^ In contrast, male fetal sex is associated with a pro-inflammatory state in the first trimester, which may be correlated to the historically high rates of early pregnancy loss among male fetuses compared to females.^10,31,32^ Steen et al., postulated that testosterone may act to protect male fetuses against preeclampsia by dampening the maternal immunological response to the male fetus as pregnancies with a male fetus have higher serum concentrations of testosterone.^17^ However, studies have demonstrated a positive correlation between serum testosterone and preeclampsia^33^, which is in accordance with our finding of increased risk in preeclampsia among female/male twin pairs compared to female/female.

Individuals carrying twin pregnancies are at an increased risk of preeclampsia compared to those carrying singleton pregnancies.^2,3,34,35^ Studies have suggested that dichorionic twins predispose to preeclampsia development due to an increase of placental mass in the womb. According to Funaki et al., there is a positive correlation between the amount of placental tissue present and maternal immune system activation.^3^ Other studies have found no differences in maternal outcomes by chorionicity.^36^ Furthermore, studies examining associations between zygosity and hypertensive disorders have been conflicting and inconclusive.^16^ Although there is limited evidence to support a placental mass dependent immune response, our results were consistent with this rationale. Female/male twin pairs are dizygotic, possessing an increased placental mass compared to monozygotic twins.^35^ Secretion of sFlt1 proportional to the large placental mass may contribute to an increased risk of preeclampsia in female/male fetal sex pairs. However, zygosities and chorionicities were not evaluated for our entire sample population, therefore we are unable to draw a definitive conclusion.

One of our study strengths includes using a diverse study population with low levels of missingness among our variables. Also, our perinatal database undergoes frequent quality checks and contains extensive information on each pregnancy occurrence. Due to this, we were able to explore several hypertensive disorders and different preeclampsia subtypes. Some of our limitations include our small sample size, a common issue in studies involving twins due to their low prevalence in the general population. Another limitation is that we could not capture early fetal loss in monochronic twin pregnancies. This is also common among twin studies, as early perinatal fetal losses frequently occur in twin deliveries due to vascular complications that arise from sharing a placenta. Lastly, we could not rule out the potential of residual confounding.

### Perspectives

Our study demonstrated an increased risk of preterm preeclampsia in female/male fetal sex pairs compared to female/female pairs. This area is understudied and there is not there is not sufficient evidence to suggest that fetal sex is a predictive marker of preeclampsia risk. The mechanisms that may explain this association have not been explored as they have in singleton pregnancies. There is a need for additional research on preeclampsia in multifetal pregnancies more generally, particularly to understand subtypes within these populations. Studies should consider fetal sex when exploring biological mechanisms driving preeclampsia in multifetal pregnancies.

## Data Availability

The data used in this manuscript was provided by Peribank under license/permission. Access to data requires permission from Peribank

## Novelty and Relevance

1. What is New?

a. We found that fetal sex pairs in twin pregnancies may influence risk of preeclampsia.
2. What is Relevant? (How the study related to hypertension)

a. Preeclampsia is a maternal hypertensive disorder that increases risk of cardiovascular disease, but multifetal pregnancies are often excluded from studies, thus identifying risk factors in this population is important.
3. Clinical/Pathophysiological Implications

a. Sexual dimorphism exists in preeclampsia risk among twin pregnancies, but mechanisms are unexplored.

## Acknowledgments, Source of Funding, & Disclosures

**Acknowledgements**

We thank Peribank for allows us access to their data and all study participants. In Peribank, subject data were obtained following full informed subject consent with generous support from the Departments of Obstetrics and Gynecology and Pathology and Laboratory Medicine at Texas Children’s Hospital and Baylor College of Medicine on the Peribank Protocol (IRB H-26364, Dr. Kjersti Aagaard PI)

## Source of Funding

This research is funded in part by the National Institutes of Allergy and Infectious Diseases (NIAID) grant 1RO1AI141501-O1A1 to B.D.T. The content is solely the responsibility of the authors and does not necessarily represent the official views of the National Institutes of Health.

## Disclosures

None

## Notes

### Competing Interest Statement

The authors have declared no competing interest.

### Author Declarations

The Institutional Review Board (IRB) at the University of Texas Medical Branch.

